# Feasibility of intravaginal artesunate as an adjuvant HPV & cervical precancer treatment among women living with HIV in Kenya: Study protocol for a phase II clinical Trial

**DOI:** 10.1101/2025.06.02.25327779

**Authors:** Annum Sadana, Jackton Omoto, Katherine Sorgi, Lisa Rahangdale, Jennifer Smith, Mihaela Plesa, Chemtai Mungo

**Author notes:** **Corresponding Author:** Chemtai Mungo, MD, MPH.

## Abstract

**Background:** Cervical cancer disproportionately affects women in low- and middle-income countries (LMICs), who account for 90% of deaths from the disease. Human papillomavirus (HPV) is responsible for 99% of cervical cancer cases. Women living with HIV (WLWH) have a higher risk of persistent HPV infection and a greater likelihood of developing cervical cancer. Prevention of cervical cancer requires effective screening and precancer treatment programs. In LMICs, the common treatment method for cervical precancer is thermal ablation. However, for WLWH, thermal ablation is associated with high rates of persistent HPV infection following treatment, a key risk factor for precancer recurrence. Adjuvant topical treatments with cytotoxic or antiviral properties may reduce HPV persistence HPV following ablation. Preclinical and early-phase clinical trials indicate that topical artesunate is active against HPV-associated anogenital lesions, including cervical precancer, and can induce HPV clearance. Consequently, intravaginal artesunate may improve HPV clearance following thermal ablation, although no clinical trials have investigated this.

**Methods:** We are conducting a phase II, double-blind, randomized, placebo-controlled trial among 120 HIV seropositive women in Kenya to investigate the feasibility of self-administered intravaginal artesunate pessaries as adjuvant therapy following thermal ablation treatment for cervical precancer. The primary outcome is type-specific HPV clearance six months after randomization. Secondary outcomes are safety, adherence, acceptability, uptake, and retention. Participants will use self-administered artesunate pessaries starting at least 4 weeks following ablation, on weeks 1,3,5 with alternating drug-free weeks, with a follow-up period extending up to 24 weeks after randomization.

**Discussion:** High rates of persistent HPV infection in WLWH is a key limitation of thermal ablation, the most accessible cervical precancer treatment in LMICs. This trial will investigate the feasibility of repurposing topical artesunate as an adjuvant therapy to improve HPV clearance following thermal ablation in WLWH.

**Trial Registration:** ClinicalTrials.gov identifier: da

## Introduction

Although cervical cancer is a preventable disease, there were 660,000 new cases and 350,000 deaths from the disease globally in 2022 ^1,2^. This burden is disproportionately shouldered by low-and middle-income countries (LMICs), such as those in Sub-Saharan Africa where cervical cancer is the leading cause of cancer deaths among women ^1^. Additionally, women living with HIV (WLWH), the majority of whom live in LMICs, have a higher incidence and persistence of high-risk human papillomavirus (hrHPV) and up to six times increased risk of developing cervical cancer ^3^. In response to the global burden of cervical cancer, the World Health Organization (WHO) launched the 90/70/90 cervical cancer elimination goals in 2018, setting targets for cervical cancer elimination ^4^. Recognizing that human papillomavirus (HPV) is the causative agent for virtually all cervical cancer ^5^, the 90/70/90 goals aim to prevent millions of cases of and deaths due to cervical cancer through HPV vaccination, screening, and treatment of precancerous lesions. Namely, at least 90% of women with cervical precancer should receive adequate treatment.

Supported by the World Health Organization (WHO) guidelines, most LMICs, including Kenya, use a ‘screen & treat’ approach where a positive screening test (either HPV or VIA) is followed by immediate treatment without pathology confirmation of the diagnosis to reduce loss to follow-up ^6^. Current treatment options for cervical precancerous lesions, including cervical intraepithelial neoplasia grades 2 and 3 (CIN2/3), include excisional procedures such as Loop Electrosurgical Excision Procedure (LEEP) and ablative methods like thermal ablation and cryotherapy. In most LMICs, thermal ablation is the most accessible and widely used cervical precancer treatment method, as it can be performed by trained nurses and uses a portable, battery-powered device ^7^ ^8^. To achieve the WHO goal of treating 90% of women who screen positive for cervical precancer ^4^, the Kenya Cancer Guidelines recommend thermal ablation for the treatment of cervical precancer among both HIV-positive and HIV-negative women as part of a screen-triage-and-treat program ^8^.

In recently published results from a non-inferiority trial conducted by the International Agency for Research on Cancer (IARC) in Zambia, treatment success (defined as either type-specific HPV clearance or a negative VIA test) was substantially lower in HIV-positive women (62.1%), irrespective of whether they underwent thermal ablation, cryotherapy, or LEEP, compared to HIV-negative (85.8%) women at 6 and 12 months follow-up ^9^. However, in a randomized clinical trial (RCT) of HIV+,hrHPV+, women with CIN2+ conducted in Kenya, WLWH who underwent cryotherapy were more likely to have persistent hrHPV infection at 12 months (61%) compared with those who underwent LEEP (49%) ^10^. In the same Kenya RCT, women with type-specific persistent hrHPV detected at 12 months were 5 times more likely to have recurrent CIN2+ at 12 months or later compared to those who did not have hrHPV (HR, 5.28; 95% CI, 2.78-10.04, p<.001 ^10^.

Among HIV-negative women, thermal ablation is highly effective for treating cervical precancer, with a 92–95% cure rate for CIN2/3 (defined by normal cytology or histology on follow-up) ^11^. In contrast, studies of thermal ablation treatment among women living with HIV (WLWH) show higher rates of CIN2/3 treatment failure and persistent HPV infection. A Kenyan study of WLWH with biopsy-confirmed CIN 2/3 reported a 66% cure rate after treatment with thermal ablation (defined as regression to CIN1 or normal histology) at 12-month follow-up ^12^. Similarly, in an Indian study of 52 WLWH with CIN2/3, of whom 32 were treated with thermal ablation, only 66% had histologic regression, and only 44% exhibited HPV clearance at 3+ years^13^. In this study, only 44% had HPV clearance at the 3-year follow-up ^13^. Many studies use HPV clearance as a marker of treatment success due to its high negative predictive value for CIN2/3 recurrence ^14–18^. Preliminary results from a Zambian non-inferiority trial of 197 women treated with thermal ablation indicate that at 6 months follow-up after thermal ablation, treatment success (defined as either type-specific HPV clearance or a negative VIA result) was 83% in HIV-negative women but only 44% in women living with HIV^19^. In an analysis of the completed non-inferiority trial, in the thermal ablation arm, WLWH had 40.1% clearance of baseline HPV, whereas HIV-negative women had 62.6% clearance (Dr. Richard Muwonge, personal communication, April 2025).

Thermal ablation destroys cervical tissue to a depth of roughly 2.6 – 3.5 mm ^20^. This may be inadequate for under-screened women in LMICs, including WLWH, whose CIN2/3 lesions may be deeper. In a Peruvian cohort, 10.3% of excised CIN2/3 specimens had a lesion depth of greater than 3.5 mm, suggesting that a 3.5 mm depth of tissue necrosis may not fully destroy HPV-infected precancerous tissue in this population ^21^. Additionally, current ablation probes may not reach HPV-infected crypts deeper in the endocervical canal, leaving these areas untreated during thermal ablation ^22,23^. The limited efficacy of thermal ablation among WLWH highlights an urgent need for strategies to improve cervical precancer treatment outcomes in this high-risk population.

Topical agents with cytotoxic or antiviral properties may selectively target residual HPV-infected cells after thermal ablation and improve HPV clearance. An ongoing randomized trial is evaluating the feasibility of topical, self-administered intravaginal 5-Fluorouracil (5FU) as an adjuvant to LEEP to reduce CIN2/3 recurrence among WLWH in South Africa (NCT0541381)^24^. If randomized efficacy trials support their safety and efficacy, low-cost and accessible self-administered topical agents such as 5FU could be repurposed as adjuvant therapies, improving the effectiveness of current cervical precancer treatments and hence strengthening secondary prevention of cervical cancer in WLWH.

Given its known safety profile, one of the proposed topical self-administered therapies is artesunate—an essential WHO medication—and its active metabolite, dihydroartemisinin (DHA)^25^. Artemisinin and its derivatives show potential in cancer treatment due to their ability to induce ferroptosis, an oxidative, iron-dep ndent form of regulated cell death (ferritin is overexpressed in many cancer cell lines) ^26^ and cytotoxicity ^27^. Artesunate and its active metabolite dihydroartemisin (DHA) exhibit selective cytotoxicity against HPV-immortalized and cervical cancer cells, with minimal toxicity to normal cervical epithelial cells ^28^. Experiments on *in vivo* canine models infected with canine HPV show that topical DHA treatment effectively inhibits tumor formation without ulceration of normal epithelium ^28^.

In 2020, Trimble et al. published data from a proof-of-concept study of intravaginal artesunate suppositories for primary treatment of CIN2/3 among HIV-negative women in the United States^29^. In this phase I, trial dose-escalation study involving 28 women with biopsy-confirmed CIN2/3, the self-administration of three five-day cycles of intravaginal 200 mg artesunate pessaries proved to be safe and well-tolerated. 19 of 28 (67.9%), subjects had histologic regression to CIN1 or less in an intention-to-treat analysis, with 9 of the 19 whose lesions underwent histologic regression (47.4%) experiencing clearance of HPV genotypes detected at baseline.

A phase II randomized placebo-controlled trial of Artesunate vaginal inserts for the treatment of CIN2/3 among HIV sero-negative women is currently enrolling subjects at several U.S. sites, with no serious adverse events reported so far (NCT04098744). Two other randomized, placebo-controlled studies of topical artesunate for the treatment of intra-anal and vulvar high-grade intraepithelial lesions (HSIL), respectively, are ongoing in the US (NCT06206564, NCT06075264). The vaginal inserts referenced in some of these trials are equivalent to pessaries used in our Phase II trial, we use this terminology interchangeably.

Although preclinical and early-phase clinical data support topical artesunate for the treatment of HPV-associated anogenital lesions, its potential as an adjuvant to improve HPV clearance post-thermal ablation in WLWH has not yet been investigated. To address this gap, we are conducting a Phase II, placebo-controlled, feasibility trial to evaluate intravaginal artesunate as an adjuvant therapy after thermal ablation in WLWH in Kenya.

## Materials and Methods

### Hypothesis and Specific Aims

We hypothesize that the use of self-administered artesunate pessaries (vaginal inserts) as adjuvant therapy following ablation among WLWH will result in improved HPV clearance at six months in the artesunate arm compared to the placebo arm.

#### Primary Objective

To estimate type-specific HPV clearance rates at 24 weeks after randomization among WLWH randomized to adjuvant self-administered artesunate vaginal inserts vs. placebo inserts following thermal ablation in Kenya.

#### Secondary Objectives

1. To evaluate the safety of adjuvant artesunate vaginal inserts vs placebo following thermal ablation among WLWH
2. To evaluate adherence, defined as the use of at least 80% (12 of 15) self-administered artesunate or placebo vaginal inserts following thermal ablation in WLWH
3. To investigate the acceptability of intravaginal self-adminstered artesunate as adjuvant therapy following thermal ablation in WLWH
4. To evaluate study uptake, accrual, and retention rates through week 24

#### Exploratory Objectives

1. To investigate longitudinal changes in the cervicovaginal microbiome (VMB) following intravaginal artesunate pessary vs. placebo use among WLWH.
2. To investigate the frequency and magnitude of genital HIV-1 shedding and measures of local immune activation following intravaginal artesunate pessary vs. placebo use among WLWH

### Study Design

This is a phase II, randomized, placebo-controlled trial.

### Study Population and Setting

This study will be conducted in two sites, both in the western Nyanza region of Kenya: Kisumu County and Homabay County. Both Kisumu and Homabay County have a population of over 1.1 million people each ^30^. The prevalence of HIV in adult females (ages 15-49) is roughly 20% in Homabay County and 19% in Kisumu County. These rates are roughly four times higher than the national HIV prevalence among females, which is 4.5% ^31,32^. There is a co-occurrence of HIV and HPV infection in these counties, with some studies indicating a rate close to 20% among a population of HIV-positive and negative women, over double the national prevalence of 9% ^33^. In a cervical cancer HPV screening trial conducted in western Kenya, among 771 HIV- positive women, 35% were HPV-positive, compared to 16% of HIV- negative women ^34^.

Kisumu has one teaching and referral hospital, five County referral hospitals, nine sub-county hospitals, over 50 dispensaries, and over 15 health centers ^35^. In Homabay County, there is 1 referral hospital, and there are eight sub-county hospitals, over 40 dispensaries, and as many health centers ^36^. LEEP is offered at Jaramogi Oginga Odinga Teaching and Referral Hospital (JOOTRH) in Kisumu and at Matata Hospital, Homabay County Referral, and Rachunyo South Hospital in Homabay County. However, several facilities in Kisumu County and Homabay County offer screening and thermal ablation services, including Lumumba Sub-County Hospital and Matata Hospital, where the studies are based. Most women who need treatment for confirmed or suspected cervical precancer in western Kenya are treated with thermal ablation as access to LEEP is limited.

### Eligibility and Participant Recruitment

Participants will be recruited from clinical facilities providing performing thermal ablation in western Kenya, including, but not limited to Kisumu, Homabay, Vihiga, Kakamega, and Siaya counties. The study team will give educational talks about the study protocol as part of community outreach activities at these sites. Women interested in participating will be screened for eligibility and subsequently enrolled and consented if all eligibility criteria are met.

Enrollment for this study will be done on a rolling basis at both study sites with a maximum enrollment of 120 women.

#### Study Procedures

WLWH who are HPV positive and treated with thermal ablation, will be recruited according to the inclusion criteria in Table 1.

**Table 1.**
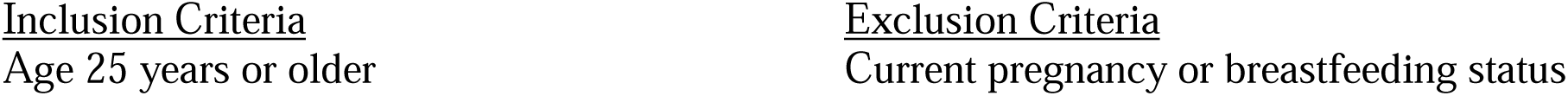
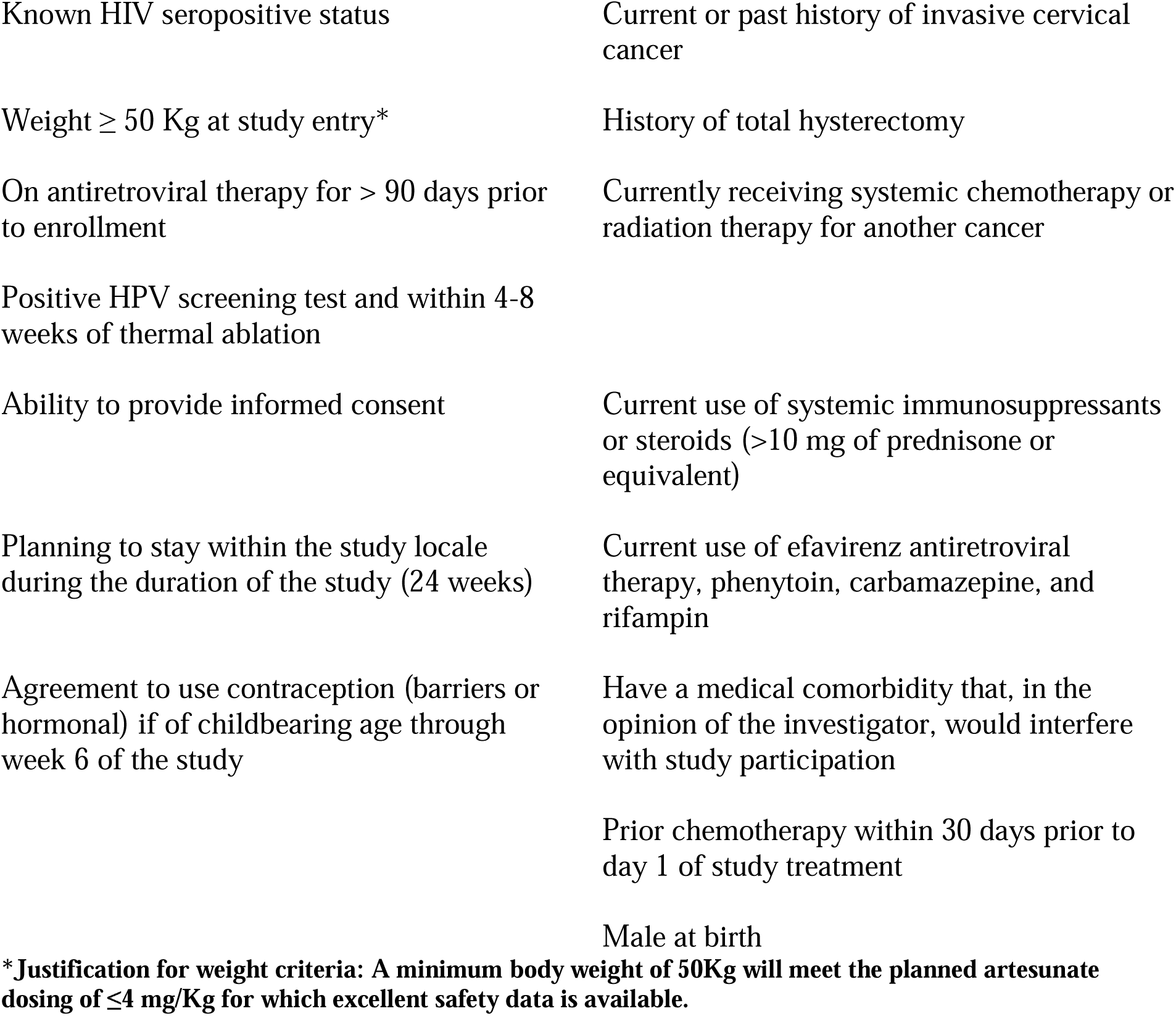
Inclusion and Exclusion Criteria.

#### Trial intervention and assignment

This is a double-blind, randomized, placebo-controlled trial. Eligible participants will be randomly assigned to either 200 mg artesunate (intervention arm) or placebo (control arm) in a 2:1 ratio. The placebo formulation will contain the same inactive ingredients as the artesunate insert without active artesunate. Placebo and artesunate pessaries have the same appearance, weight, and packaging. An unblinded statistician not involved in the trial will prepare the randomization sequence. The unblinded study pharmacist will create blinded cartons per the randomization sequence with three dosing cycles each. At the time of randomization, a kit number will be generated electronically. The site pharmacist, who does not interact with study participants, will select the corresponding carton, labeled with only the randomization number. Study staff and participants will be blinded to the treatment arm. The study procedures and subsequent follow-up visits are outlined in Figure 1.

**Figure 1.**
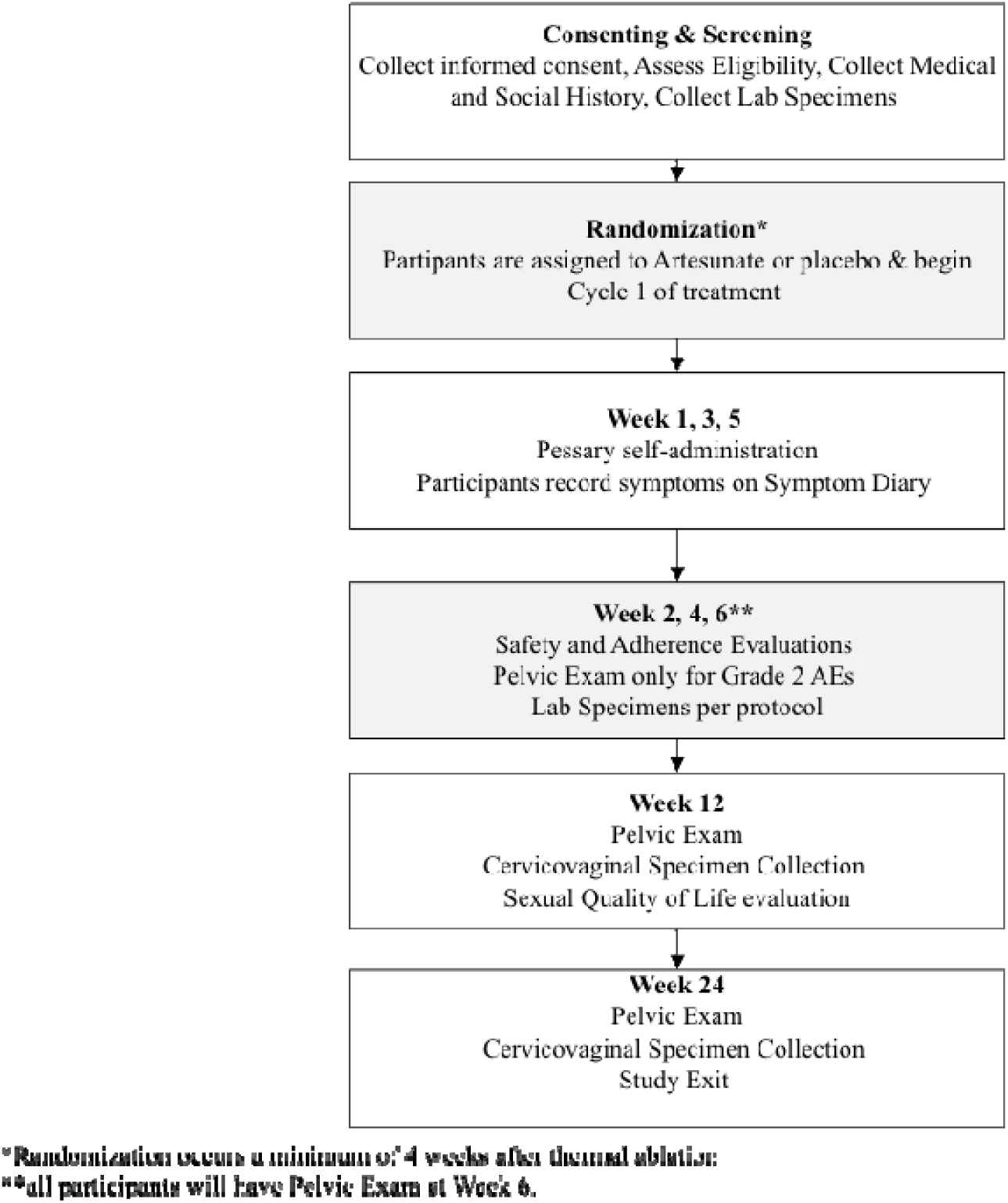
Study Flow Diagram.

#### Study Procedures by Visit

##### Screening Visits

Eligible participants will complete a written informed consent. Illiterate participants will be consented with a witness present and provide a thumbprint for consent. A comprehension checklist will be utilized to ensure adequate comprehension of all study activities. Participants without a pre-ablation HPV test will undergo HPV testing via Atila ScreenFire HPV assay as part of the screening activities. Baseline demographic and clinical data will be collected, including sexual, reproductive history, and HIV diagnosis and treatment history. A trained nurse will collect baseline samples including blood samples for HIV, CD4, and malaria antigen testing. Participants who test positive for malaria with a Rapid Detection Test (RDT) will be offered treatment per the local standard of care before enrollment. A study nurse will perform a pelvic exam to evaluate for post-thermal ablation healing and to evaluate any other abnormalities; findings will be recorded. Cervicovaginal swabs (CVS) will then be collected including those for HPV genotyping, microbiome analysis, DNA methylation, inflammatory markers, and sexually transmitted diseases (STIs) testing (Table 2). Participants will receive treatment for *Neisseria gonorrhoeae*, *Chlamydia trachomatis*, and *Trichomonas vaginalis* according to the results of the screening tests. All other CVS samples will be stored. Screening procedures can be completed up to 5 weeks following consenting.

**Table 2.** Cervicovaginal Samples.

| **Cervicovaginal swabs to be collected (Screening, Week 6, Week 12, Week 24)** |
| --- |
| 1. HPV Genotyping |
| 2. Microbiome |
| 3. Methylation |
| 4. Inflammatory markers |
| 5. Sexually transmitted infections (STIs) |

##### Randomization/Enrollment

Participants will be enrolled and randomized to either the artesunate or placebo arm during Week 1, when artesunate or placebo dosing begins. At this visit, all participants receive comprehensive education on all study procedures, including self-administration of the pessaries using a vaginal applicator, both through pictorial aids (see Appendix 1) and a pelvic demonstration model, information about biweekly study visits during the dosing phase, guidance on maintaining a symptom diary, and planned weekly reminder phone calls and text messages to assess adherence and adverse events. Literate participants will record any adverse events (AEs) in a symptom diary provided. For illiterate participants, AEs will be assessed verbally and on clinician assessments during the safety visits described below. Following randomization, participants will have the option to apply the first vaginal insert in the study clinic to ensure proper use and provide an opportunity for teaching reinforcement. Participants will be provided the four remaining vaginal inserts for the first cycle, four vaginal applicators, and four tampons to continue daily use for the remainder of week one. At this visit, the participant will also have a survey administered by a study research assistant to evaluate baseline Sexual Quality of Life (SQoL).

##### Weeks 1, 3, 5

Weeks 1, 3, and 5 mark the three dosing weeks of the study. During these weeks, participants will self-administer a vaginal pessary nightly for five consecutive days. Following the application of the pessary using a study-provided applicator, participants will be encouraged to insert a tampon overnight to keep the pessary at the cervix. Following nightly application of the pessary and tampon, participants will be instructed to place the used applicators, as well as the pessary packaging, in a resealable bag to return to the clinic at the next visit. They will be instructed to remove the tampon the following morning. To ensure participant safety, non-superabsorbent tampons will be provided, with clear instructions to not keep them in for longer than 10 hours to minimize the risk of toxic shock syndrome. Participants will be instructed to abstain from sexual intercourse on dosing days, after applying the pessary, to minimize irritation; however, sexual intercourse before pessary insertion will not be prohibited. Participants will self-monitor for AEs as described above. Weekly phone calls from study staff will serve to review usage instructions, document adverse events (AEs), and address participant questions. If menstruation occurs during weeks 1, 3, or 5, use of the pessary will be delayed until the end of the menstrual cycle, at which time the dosing cycle will be resumed.

##### Safety Visits (Week 2, 4, and 6)

Weeks 2 and 4 are off weeks of artesunate treatment. Participants will return to the clinic on these weeks with their used applicators, pessary covers, symptom diary, and study calendar. Study staff will review participants’ symptom diaries and study calendars and assess the frequency and severity of adverse events using the Division of AIDS (DAIDS) Adverse Event Grading Tables for Use in Microbicide Studies ^37^ and the U.S National Cancer Institute Common Terminology Criteria for Adverse Events, v5.0 (CTCAE 5.0) ^38^. Participants reporting grade 2 or worse pelvic or genitourinary AEs will undergo a pelvic examination to evaluate for any AEs, such as inflammation or ulceration. For adherence assessments, used applicators and pessary packaging will be collected at each visit. Adherence will be reinforced by reviewing the artesunate administration checklist and a comprehension checklist at each safety visit. Participants will then receive the next set of five vaginal inserts, applicators, and tampons. Week 6 marks the last safety and adherence evaluation visit. During this visit, all participants will receive a pelvic exam to evaluate cervicovaginal tissues for inflammation after the final treatment cycle. CVS samples will be collected (same samples as Screening). Participants will take another standardized assessment for SqoL.

##### Week 12, 24

At weeks 12 and 24, participants will return for cervicovaginal sample collection as described in Table 2. Week 24 will mark the last study visit, after which participants will continue with standard of care follow-up including yearly screening practices for cervical precancer.

### Storing and processing of biospecimen samples

#### 1. At baseline

Malaria rapid detection tests will be conducted on-site. HIV and CD4 tests will be shipped to a licensed lab per site protocols for time and temperature.

STI specimens (*N. gonorrhoeae, C. trachomatis, and T. vaginalis*) at screening will be processed via Gene Xpert (NG/CT) or OSOM rapid test kits in either Homabay or Kisumu. Other CVS samples will be stored as described below.

#### All other visits

All specimens at other visits will be logged and stored at the Universities of Nairobi, Illinois, and Manitoba Research and Training Centre (UNIM) laboratory, Kisumu or VIBRI laboratory, Homabay. Conditions of storage and processing will follow the manufacturer’s directions for specific assays. Vaginal swabs will be immediately stored in a sterile sample collection tube post-collection. The laboratory technician will ensure that tubes are appropriately pre-labeled with the PID and date of collection before sample collection events. Specimen shipping will use a courier that maintains cold-chain transport when needed. HPV genotyping will occur at the University of Manitoba.

#### Data Management

All data will be linked to a participant identification number (PID) and de-identified to minimize risks of disclosure. Data will be stored in the UNC REDcap database and analyzed using STATA version 14.0. Identifiers and all locator forms will be kept in a locked cabinet at each study site, and electronic copies will be directly stored on password-protected encrypted cloud servers and only accessed by authorized individuals.

#### Sample Size considerations

The proposed sample size is based on the primary objective, which is to compare the rate of type-specific hrHPV clearance 24 weeks (six months) after thermal ablation by randomization arm. Type-specific hrHPV clearance was chosen as a proxy clinical outcome, given the known relationship between persistent hrHPV infection and cervical precancer recurrence following treatment ^12^ ^10^. Current best estimates of type-specific hrHPV clearance at 24 weeks following thermal ablation among WLWH come from a study in Zambia that reported a 40-44% rate of type-specific hrHPV clearance in this population following thermal ablation ^9,19^. This estimate is assumed to approximate the HPV clearance rate we expect to observe in the placebo arm of our Phase II RCT. In the single-arm phase I trial of Artesunate for primary treatment of CIN2/3 among HIV-negative women in the U.S., ^29^ 47.9% (9/19) had clearance of HPV genotype detected at baseline at the Week 42 follow-up. We use these data to estimate the effect of artesunate as adjuvant therapy for this trial, as no studies directly evaluate the effect size of adjuvant artesunate on HPV clearance in WLWH. Given the known lower rates of hrHPV clearance following treatment in WLWH compared to HIV-negative women, ^39,40^ we conservatively estimate a 25% effect size of artesunate for this planned trial. Under this assumption, participants in the treatment arm would have 69% hrHPV clearance at six months, compared to 40-44% in the placebo arm, as observed in the 2020 trial and 2024 results from the trial in Zambia. Using a 2:1 randomization (artesunate: placebo), a total of 120 participants will be randomized, with 80 in the artesunate arm and 40 in the placebo arm to achieve 80% power to detect a difference of 25% between arms. The test statistic is based on a t-test approximation to binomial with a one-sided alpha = 0.05, and the sample size is adjusted for a 10% loss to follow-up in each arm.

### Outcomes & Analysis Plan

#### Primary Objective

Estimate type-specific high-risk HPV clearance rates at 24 weeks among WLWH randomized to adjuvant self-administered Artesunate vs. placebo vaginal inserts following thermal ablation

<u>Primary Endpoint:</u> Clearance of type-specific high-risk HPV genotype(s) at 24 weeks present at baseline.

<u>Statistical Analysis:</u> The number and proportion of women in each study arm who demonstrate clearance of the specific hrHPV genotype, or genotypes present at baseline, at six months will be reported, with corresponding 95% CIs. The effect of artesunate on hrHPV clearance will be estimated by the difference in these proportions between the trial arms, with a corresponding 95% CI, and tested using a 2-sample normal approximation to the binomial distribution. A complete case intention-to-treat (ITT) analysis will be conducted, with multiple imputation methods for missing data as sensitivity analyses.

#### Secondary Endpoints

##### 1. Safety

<u>Endpoint</u> Type, frequency, severity and duration of adverse events (AEs) using NCI CTCAE v5.0 and DAIDS standardized scales.

<u>Statistical Analysis:</u> The counts of adverse events for each participant in both HIV status groups will be tabulated by severity (grades 1-5). The proportion of participants with a severe AE (grade 3 or higher) within each treatment group will be reported along the exact (Clopper-Pearson) one-sided upper 95% confidence bounds. The proportion of participants who report a severe AE (if any) will also be reported, along with an exact one-sided upper 95% confidence bound.

##### 2. Adherence

<u>Endpoint</u>: Confirmed self-administration of at least 80% (12 of 15) vaginal inserts (yes/no).

<u>Statistical Analysis</u>: Two sources of adherence data (self-report and counts of returned packaging for used pessaries) will be collected, and corresponding adherence rates will be reported. Using mixed effects models to account for repeated longitudinal measurements per woman, the intra-class correlation coefficient (ICC) will be estimated to quantify within-woman agreement between the three adherence measurement methods ^41^. Assuming high agreement (correlation of 75% or higher), a primary composite adherence rate will be computed by averaging the data from both adherence assessment methods (each carrying equal weight). A participant will be classified as adherent if she administers at least 12 of 15 (80%) of vaginal inserts. We will report the number and percent of women achieving adherence with the corresponding 95% CI for each study arm. The endpoint will be compared between study arms using an estimated difference in proportions with a corresponding 95% CI and a Fisher’s exact test.

##### 3. Acceptability

<u>Endpoint:</u> Response to a close-ended structured acceptability questionnaire at Week 6 <u>Statistical Analysis</u>: Responses to an acceptability questionnaire will be summarized, including means and standard deviation for responses to questions graded on a Likert scale and proportions and 95% confidence intervals for yes/no questions. Impact of artesunate pessary used on Sexual Quality of Life will also be summarized as part of acceptability analyses, including means and and standard deviation for categories of SQoL and percentage of participants with sexual dysfunction.

##### 4. To evaluate study uptake, accrual, and retention rates through week 24

<u>Uptake endpoint:</u> Number of screen-eligible participants who agree to enroll in the study. <u>Statistical Analysis</u>: We will report the proportion of screen-eligible participants who agree to participate in the study with a corresponding 95% confidence interval.

<u>Accrual rate endpoint:</u> Number of participants that enrolled in the trial divided by time from first enrollment to last enrollment in months.

<u>Statistical Analysis:</u> We will report the time from the first accrual to the last accrual, divided by the number of participants enrolled in the trial.

<u>Retention rate endpoint:</u> Number of women in each study arm retained in the study over the 24-week study duration.

<u>Statistical Analysis:</u> We will report the proportion of participants retained in each study arm at each timepoint as well as through week 24 (i.e., those who attend week 24 study visit), with a corresponding 95% confidence interval. 24-week retention will be compared in the artesunate vs placebo arms using an estimated difference in proportions and corresponding 95% CI. Kaplan-Meier curves by study arm will be used to describe weeks from randomization to the last attended study visit with follow-up censored at the date of last study contact.

**Exploratory Outcomes** will include longitudinal change in the cervicovaginal microbiome (CVM), the frequency and magnitude of genital HIV-1 shedding, and measures of local immune activation over the study period. Cervicovaginal specimens will be collected and stored for future analysis, including measurement of changes in HIV-1 genital shedding and in the expression of biomarkers of local immune activation such as IFN-α2, IFN-Υ, IL-10, IL-12, IL-1α, TNF, CD8 (effector T cells), CD71 (transferrin receptor), and cleaved caspase 3 (apoptotic cell death).

Changes in the cervical microbiome will be assessed using several techniques, including evaluating the diversity of the bacterial taxa by identifying community state types (CSTs) and evaluating changes in the Lactobacillus-dominant environment.

## Discussion

In LMICs, thermal ablation is widely used to treat cervical pre-cancer, both due to its lower cost and ease of implementation. However, WLWH experience high rates of HPV persistence even after treatment and are therefore at high risk of cervical precancer recurrence and future cervical cancer incidence. Topical cytotoxic and antiviral agents have the potential to lower HPV persistence and prevent progression of cervical precancer. This phase II, randomized, placebo-controlled trial will provide preliminary data on the impact of artesunate pessaries as an adjuvant treatment for thermal ablation on hrHPV clearance among WLWH. U.S.-based trials have demonstrated the potential efficacy of Artesunate intravaginal treatment in hrHPV clearance. Therefore, we anticipate that artesunate, compared to placebo, will increase the clearance of hrHPV following thermal ablation among WLWH.

Along with our phase II trial, to strengthen the evidence base on the safety, efficacy, and feasibility of intravaginal artesunate treatment for cervical precancer, our team is currently enrolling participants in a phase I trial in Kenya (NCT06165614) of intravaginal Artesunate as a primary treatment for CIN2/3 among both HIV-negative and HIV-positive women. This trial investigates how contextual and social factors in Kenya may impact the acceptability and adherence to this intervention, as well as safety among HIV-positive women. Additionally, we recently completed a phase 1 study to investigate the pharmacokinetic properties of intravaginal artesunate (NCT06263582), which is critical in understanding the safety of intravaginal artesunate in areas with high malaria prevalence ^42(p1)^.

## Current Status

The study opened for enrollment in December 2024.

## Trial Registration

The trial is registered under the U.S. Clinical Trial Registry (NCT06519994)

## List of Abbreviations

### Ethics Statement

This clinical trial has ethics review board approval from the University of North Carolina Chapel Hill and Amref Health Africa. Written informed consent will be obtained from all study participants.

### Funding

The author(s) disclosed receipt of the following financial support for the research, authorship and/or publication of this article: This work was funded by [details omitted for double-anonymized peer review

### Authors’ contributions

CM, conceived and designed the study, designed the study protocol, providing subject matter expertise, and revised the manuscript draft. AS oversaw study implementation, training of study staff, and prepared the manuscript draft. JO contributed to protocol implementation, providing in-country expertise and revised the manuscript draft. LR and JSS provided subject matter expertise and revised the manuscript draft. KS and MP revised the manuscript draft. All authors, in their respective roles, contributed to the study and manuscript preparation and have collectively approved the final manuscript.

## Data Availability

All data produced in the present study are available upon reasonable request to the authors

## Acknowledgments

We thank the KEMRI staff and leadership for their support in the implementation of this study. Artesunate and placebo vaginal suppositories (pessaries) were provided as in-kind support by Frantz Viral Therapeutics (Mentor, OH).

## References

1. Bray F, Laversanne M, Sung H, et al. Global cancer statistics 2022: GLOBOCAN estimates of incidence and mortality worldwide for 36 cancers in 185 countries. CA Cancer J Clin. 2024;74(3):229–263. doi:10.3322/caac.21834

2. Sung H, Ferlay J, Siegel RL, et al. Global Cancer Statistics 2020: GLOBOCAN Estimates of Incidence and Mortality Worldwide for 36 Cancers in 185 Countries. CA Cancer J Clin. 2021;71(3):209–249. doi:10.3322/caac.21660

3. Castle PE, Einstein MH, Sahasrabuddhe VV. Cervical cancer prevention and control in women living with human immunodeficiency virus. CA Cancer J Clin. 2021;71(6):505–526. doi:10.3322/caac.21696

4. Global Strategy to Accelerate the Elimination of Cervical Cancer As a Public Health Problem. 1st ed. World Health Organization; 2020.

5. Schiffman M, Doorbar J, Wentzensen N, et al. Carcinogenic human papillomavirus infection. Nat Rev Dis Primer. 2016;2(1):1–20. doi:10.1038/nrdp.2016.86

6. World Health Organization and Special Programme of Research, Development, and Research Training in Human Reproduction (World Health Organization) - 2021 - WHO guideline for screening and treatment of cervi.pdf. Accessed January 31, 2025. https://iris.who.int/bitstream/handle/10665/342365/9789240030824-eng.pdf?sequence=1&isAllowed=y

7. Castle PE, Murokora D, Perez C, Alvarez M, Quek SC, Campbell C. Treatment of cervical intraepithelial lesions. Int J Gynecol Obstet. 2017;138(S1):20–25. doi:10.1002/ijgo.12191

8. Kenya Ministry of Health. National Cancer Screening and Early Diagnosis Guidelines. Published online 2024.

9. Basu P, Mwanahamuntu M, Pinder LF, et al. A portable thermal ablation device for cervical cancer prevention in a screen-and-treat setting: a randomized, noninferiority trial. Nat Med. 2024;30(9):2596–2604. doi:10.1038/s41591-024-03080-w

10. Chung MH, De Vuyst H, Greene SA, et al. Human Papillomavirus Persistence and Association With Recurrent Cervical Intraepithelial Neoplasia After Cryotherapy vs Loop Electrosurgical Excision Procedure Among HIV-Positive Women: A Secondary Analysis of a Randomized Clinical Trial. JAMA Oncol. 2021;7(10):1514. doi:10.1001/jamaoncol.2021.2683

11. Randall TC, Sauvaget C, Muwonge R, Trimble EL, Jeronimo J. Worthy of further consideration: An updated meta-analysis to address the feasibility, acceptability, safety and efficacy of thermal ablation in the treatment of cervical cancer precursor lesions. Prev Med. 2019;118:81–91. doi:10.1016/j.ypmed.2018.10.006

12. Mungo C, Osongo CO, Ambaka J, Omoto J, Cohen CR. Efficacy of thermal ablation for treatment of biopsy confirmed high grade cervical precancer among women living with HIV in Kenya. Int J Cancer. 2023;153(12):1971–1977. doi:10.1002/ijc.34737

13. Joshi S, Muwonge R, Kulkarni V, et al. Can we increase the cervical cancer screening interval with an HPV test for women living with HIV? Results of a cohort study from Maharashtra, India. Int J Cancer. 2023;152(2):249–258. doi:10.1002/ijc.34221

14. Perkins RB, Guido RS, Castle PE, et al. 2019 ASCCP Risk-Based Management Consensus Guidelines for Abnormal Cervical Cancer Screening Tests and Cancer Precursors. J Low Genit Tract Dis. 2020;24(2):102–131. doi:10.1097/LGT.0000000000000525

15. Cuschieri K, Bhatia R, Cruickshank M, Hillemanns P, Arbyn M. HPV testing in the context of post-treatment follow up (test of cure). J Clin Virol. 2016;76:S56–S61. doi:10.1016/j.jcv.2015.10.008

16. Lin Y, Long Y, He J, Yi Q. The residual rate of HPV and the recurrence rate of CIN after LEEP with negative margins: A meta-analysis. PLOS ONE. 2024;19(3):e0298520. doi:10.1371/journal.pone.0298520

17. Inturrisi F, Rozendaal L, Veldhuijzen NJ, Heideman DAM, Meijer CJLM, Berkhof J. Risk of cervical precancer among HPV–negative women in the Netherlands and its association with previous HPV and cytology results: A follow-up analysis of a randomized screening study. PLOS Med. 2022;19(10):e1004115. doi:10.1371/journal.pmed.1004115

18. Gottschlich A, van Niekerk D, Smith LW, et al. Assessing 10-Year Safety of a Single Negative HPV Test for Cervical Cancer Screening: Evidence from FOCAL-DECADE Cohort. Cancer Epidemiol Biomarkers Prev. 2021;30(1):22–29. doi:10.1158/1055-9965.EPI-20-1177

19. Pinder LF, Parham GP, Basu P, et al. Thermal ablation versus cryotherapy or loop excision to treat women positive for cervical precancer on visual inspection with acetic acid test: pilot phase of a randomised controlled trial. Lancet Oncol. 2020;21(1):175–184. doi:10.1016/S1470-2045(19)30635-7

20. Haddad N, Hussein I, Blessing K, Kerr-Wilson R, Smart G. Tissue Destruction Following Cold Coagulation of the Cervix. J Gynecol Surg. 1988;4(1):23–27. doi:10.1089/gyn.1988.4.23

21. Taxa L, Jeronimo J, Alvarez M, et al. Depth of Cervical Intraepithelial Neoplasia Grade 3 in Peruvian Women: Implications for Therapeutic Depth of Necrosis. J Low Genit Tract Dis. 2018;22(1):27–30. doi:10.1097/LGT.0000000000000355

22. Aiyenuro A, Griffin H, Schichl K, et al. Role of Reserve Cells in Metaplasia and the Development of Human Papillomavirus-Associated High-Grade Squamous Intraepithelial Lesions at the Cervical Transformation Zone. Lab Investig J Tech Methods Pathol. 2025;105(7):104166. doi:10.1016/j.labinv.2025.104166

23. Papoutsis D, Underwood M, Williams J, Parry-Smith W, Panikkar J. Expansile Endocervical Crypt Involvement by CIN2 - 3 as a Risk Factor for High Grade Cytology Recurrence After Cold Coagulation Cervical Treatment. Geburtshilfe Frauenheilkd. 2020;80(9):941–948. doi:10.1055/a-1202-2157

24. UNC Lineberger Comprehensive Cancer Center. Acceptability and Feasibility of Combination Treatment for Cervical Precancer Among South African Women Living With HIV. clinicaltrials.gov; 2024. Accessed March 27, 2025. https://clinicaltrials.gov/study/NCT05413811

25. Ellis T, Eze E, Raimi-Abraham BT. Malaria and Cancer: a critical review on the established associations and new perspectives. Infect Agent Cancer. 2021;16(1):33. doi:10.1186/s13027-021-00370-7

26. Ma Z, Woon CYN, Liu CG, et al. Repurposing Artemisinin and its Derivatives as Anticancer Drugs: A Chance or Challenge? Front Pharmacol. 2021;12:828856. doi:10.3389/fphar.2021.828856

27. Luo J, Zhu W, Tang Y, et al. Artemisinin derivative artesunate induces radiosensitivity in cervical cancer cells in vitro and in vivo. Radiat Oncol. 2014;9(1):84. doi:10.1186/1748-717X-9-84

28. Disbrow GL, Baege AC, Kierpiec KA, et al. Dihydroartemisinin Is Cytotoxic to Papillomavirus-Expressing Epithelial Cells *In vitro* and *In vivo*. Cancer Res. 2005;65(23):10854–10861. doi:10.1158/0008-5472.CAN-05-1216

29. Trimble CL, Levinson K, Maldonado L, et al. A first-in-human proof-of-concept trial of intravaginal artesunate to treat cervical intraepithelial neoplasia 2/3 (CIN2/3). Gynecol Oncol. 2020;157(1):188–194. doi:10.1016/j.ygyno.2019.12.035

30. Kenya National Bureau of Statistics, ed. 2019 Kenya Population and Housing Census. Kenya National Bureau of Statistics; 2019.

31. National Syndemic Diseases Control Council. HIV 10 Year Progress Report.; 2024.

32. National Syndemic Diseases Control Council. Kenya HIV Estimates Portal. Published online 2025.

33. Kenya: Human Papillomavirus and Related Cancers, Fact Sheet 2023. Fact Sheet. Published online 2023.

34. Huchko MJ, Ibrahim S, Blat C, et al. Cervical cancer screening through human papillomavirus testing in community health campaigns versus health facilities in rural western Kenya. Int J Gynecol Obstet. 2018;141(1):63–69. doi:10.1002/ijgo.12415

35. KMHFR | Facilities. Accessed April 11, 2025. https://kmhfr.health.go.ke/public/facilities?page=6

36. Health Facilities Layered Under Sub-county. Homabay Health. Accessed April 11, 2025. https://homabayhealth.org/partners/health-facilities-layered-under-sub-county/

37. NIAID NIH. Female Genital Grading Table for Use in Microbicide Studies version 1.0. Published online November 2007.

38. Common Terminology Criteria for Adverse Events (CTCAE). Published online 2017.

39. Li Z, Winer RL, Ba S, et al. Effect of Human Immunodeficiency Virus Infection on Human Papillomavirus Clearance Among Women in Senegal, West Africa. J Infect Dis. 2023;227(9):1088–1096. doi:10.1093/infdis/jiac428

40. Swai P, Rasch V, Linde DS, et al. Persistence and risk factors of high-risk human papillomavirus infection among HIV positive and HIV negative tanzanian women: a cohort study. Infect Agent Cancer. 2022;17(1):26. doi:10.1186/s13027-022-00442-2

41. Cassidy CM, Rabinovitch M, Schmitz N, Joober R, Malla A. A Comparison Study of Multiple Measures of Adherence to Antipsychotic Medication in First-Episode Psychosis. J Clin Psychopharmacol. 2010;30(1):64. doi:10.1097/JCP.0b013e3181ca03df

42. Mungo C, Sorgi K, Ogollah C, et al. Phase I study on the pharmacokinetics of intravaginal, self-administered artesunate vaginal pessaries among women in Kenya. medRxiv. Published online August 12, 2024:2024.07.08.24309596. doi:10.1101/2024.07.08.24309596

