## Supplemental Material #1 for "Feasibility of intravaginal artesunate as an adjuvant HPV & cervical precancer treatment among women living with HIV in Kenya: Study protocol for a phase II clinical Trial"

### English

#### How to Use Artesunate Pessary

1

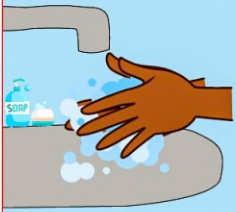


Wash your hands

2

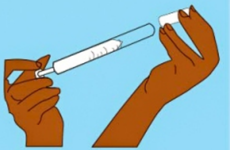

Insert the pessary into the applicator

3

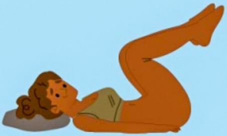

Lie with your knees drawn towards you

4

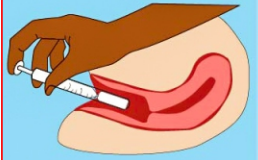

Insert the applicator into your Vagina and press the applicator plunger downward to release the pessary

5

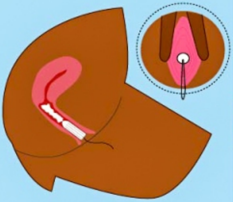

Remove the applicator and place a tampon into your vagina to prevent the pessary from coming out

6

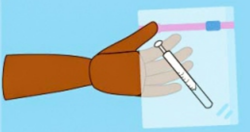

Place the used applicator into a plastic bag to return to the study clinic. Wash your hands with soap and water
